# An agent based modelling approach to study lockdown efficacy for infectious disease spreads

**DOI:** 10.1101/2020.06.22.20137828

**Authors:** Anagh Pathak, Varun Madan Mohan, Arpan Banerjee

## Abstract

We sought to simulate lockdown scenarios using an Agent Based Modelling (ABM) strategy, which is a new modelling paradigm that seeks to simulate the actions and interactions of autonomous agents within an environment. The spread of infectious viral diseases occur over a connected social network. Specifically, the goal was to understand the effect of network topology and lockdown strategies on disease spreading dynamics. To explore the effect of topology we assumed the social network over which the disease spreads to have small-world or scale-free properties characterized by a rewiring probability and degree distribution respectively. Lockdowns were simulated as intervention strategies that modified the spreading dynamics of infection over a given graph structure through changes in properties of agent interaction. Lockdown efficacy was assessed by the maximum number of infections recorded during a simulation run. Thereafter, lockdown efficacy was evaluated as a function of lockdown start times and duration. Thus, we propose that ABM approach can be used to assess various lockdown strategies that aim to prevent breakdown of medical infrastructure while accounting for realistic social network configurations specific to a local population.

**Notation:** 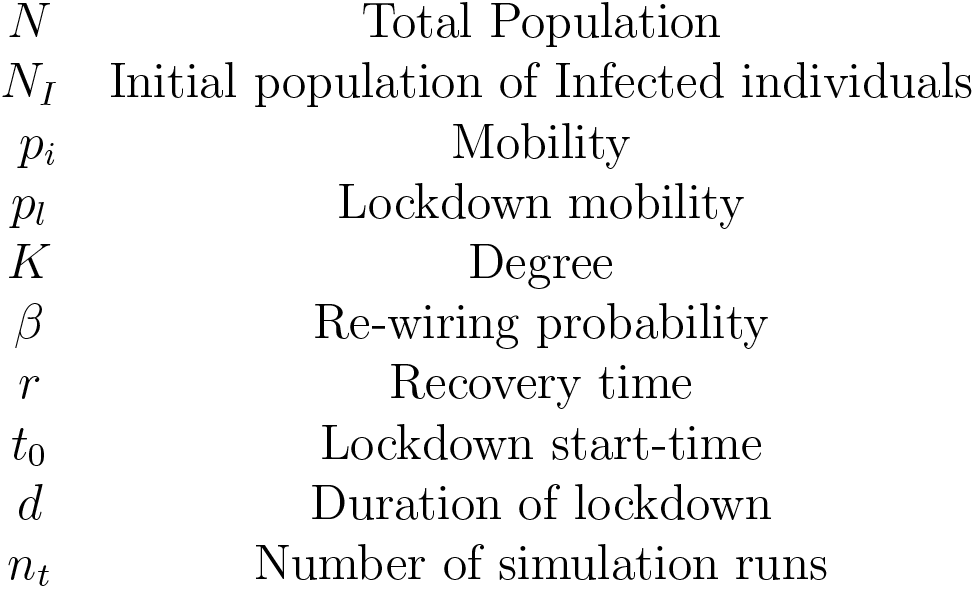

## Introduction

Understanding the dynamics of disease spread has assumed critical importance in the midst of the on-going *SARS-Cov2* pandemic. Policy-makers around the world are grappling with the difficult task of balancing the necessity of lockdown interventions to mitigate disease spread against the economic fallout of protracted disruptions to trade and commerce [Moser and Yared, 2020, Singh and Adhikari, 2020]. This problem is of critical importance in developing countries like India, where policy-makers have to tow the fine line between imposing strict lockdowns which inevitably lead to wide-spread financial hardships to economically weaker sections of the society or confront the equally grim possibility of over-straining the medical infrastructure. Therefore, modelling strategies that inform decision-making about the optimum timing and duration of lockdown interventions would be of utmost utility. However, in order to be effective, it is desirable for such models to possess relevant features of real world interactions and topologies which make it possible for real world contingencies to be mapped onto model parameters [Farmer and Foley, 2009]. Essentially a deeper understanding about the spatial aspects generated from social interactions and the temporal aspects from evolution of infection spreads requires a framing of the epidemiological questions in context of spatiotemporal network dynamics.

One relatively new paradigm that seeks to incorporate spatial and interactive features of the simulation environment is the agent based modeling (ABM) approach. Inspired by Ising and cellular automata models, ABMs regard the system as composed of agents (such as infection spread rates, mobility of population) which interact with each other based on well-defined rules [Crooks and Heppenstall, 2012, Bonabeau, 2002]. ABMs allow the introduction of environmental and interactive detailing which is not possible in ordinary differential equation based models such as the **SIR** model and its derivatives [Hethcote, 2000]. For example, with ABMs it is possible to constrain the dynamics to specific graphical topologies such as small-world or scale-free networks, that are often used to model realistic environments such as social networks or cities [Eubank et al., 2004]. In the ABM framework, it is possible to introduce detail at the scale of individual agents, making agent specific interventions almost effortless to implement, when compared to an **ODE** based model [Newman, 2018]. Over the past few years, ABMs have been demonstrated to successfully model a wide variety of realistic systems such as forest ecosystems, financial markets, ant colonies etc. [Bonabeau, 2002, Tisue and Wilensky, 2004].

The objective of this paper is two-fold. First we use an ABM approach to model disease spread while constraining the dynamics to evolve on small-world or scale-free topologies, characterized by rewiring probability and degree distribution respectively. Next, we simulate lockdown interventions by reducing the rate of interaction or mobility of infected agents in the population. In order to gauge the effectiveness of a lockdown we define the maximum fraction of infected individuals (MFII) as the maximum fraction (global) of active cases throughout the course of the epidemic. Ideally, this fraction should remain below a defined threshold; a breach of the threshold would imply that more cases exist in the population than can be handled with the available medical infrastructure. We quantify lockdown efficacy using MFII. Specifically, we seek to identify optimal temporal windows where lockdown interventions are most effective, while also studying the interaction of network topologies with lockdown severity.

## Method

### Model Description

We employ an ABM approach to study an epidemic on a static undirected **binary network** where nodes can interact only with their immediate neighbors. The simulation time-step is assumed to be 1 day for all the simulations.

The network topologies in focus are a Watts-Strogatz small-world network and a scale-free network [Watts and Strogatz, 1998, Albert and Barabási, 2002]. The small-world network was built from a ring network consisting of N nodes and **2*K=50** nearest neighbour edges, with a rewiring probability of *β*. The scale-free networks were constructed using the Barabasi-Albert preferential attachment algorithm [George, 2006]. The total number of nodes and edges were kept comparable across the two topologies for sake of valid comparison.

Initially, the disease is assumed to spread on the above mentioned topology according to the following rules: The total population for a given run is *N*. At the start of the simulation, *N*_*I*_ nodes are selected at random and designated as *‘infected’*. All other nodes are assumed to be *‘susceptible’* (**Figure 1**). At each time-step, a single neighbour of an infected node is randomly selected and designated as *‘infected’* with an interaction probability (*p*_*i*_). Once infected, a node is capable of infecting any of its susceptible neighbours, till it recovers. A running counter is maintained throughout the simulation which tracks the number of time steps since infection for all infected nodes. Infected nodes are said to *‘recover’* after a given number of time steps have elapsed (*r*: recovery time), at which point the node ceases to be infectious or capable of getting infected.

**Figure 1.**
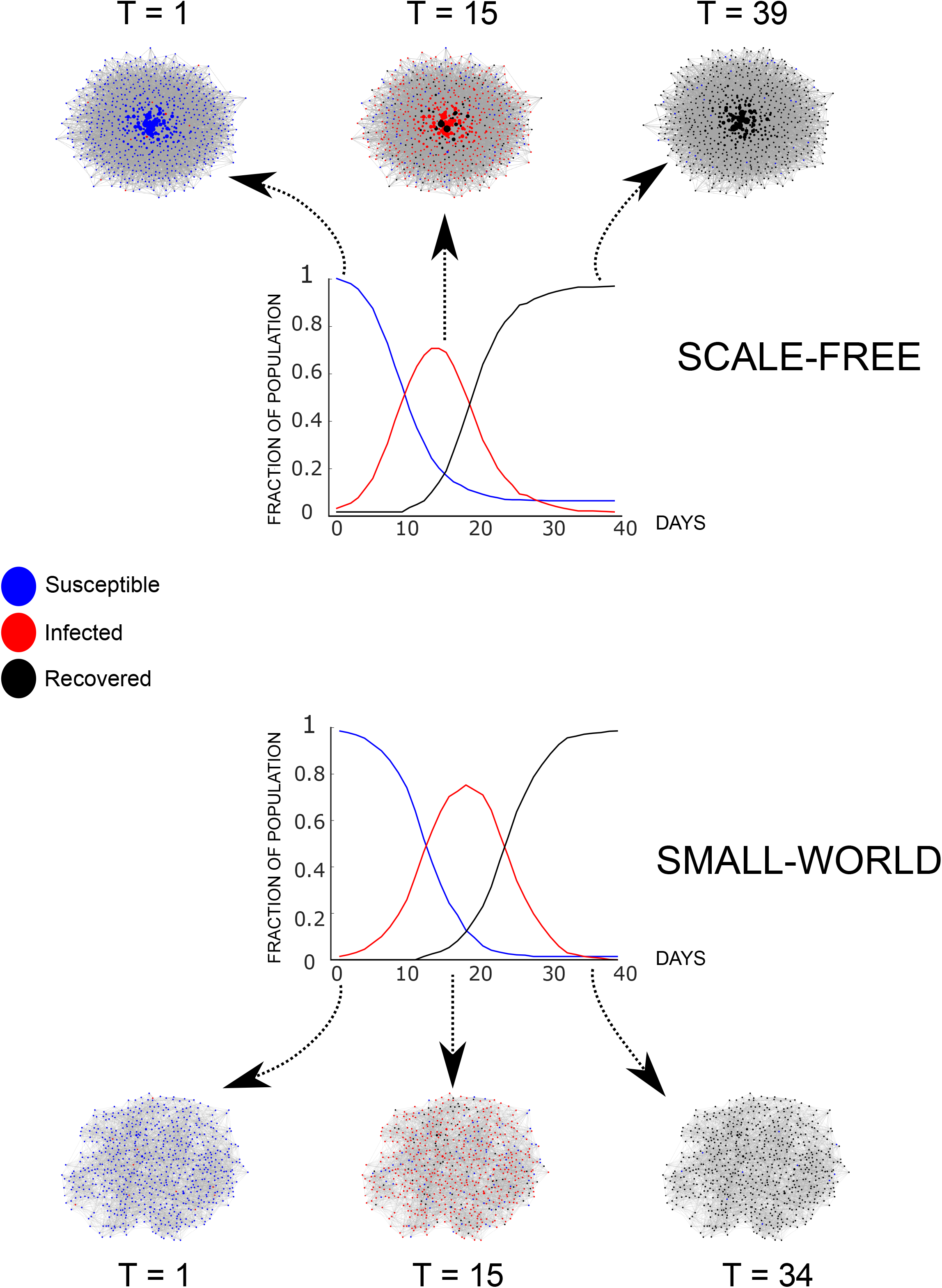
Agent based SIR model. Disease spread on **a)** Scale-free b) Small-world networks, with *N* = 700 and 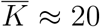; **c)** and **d)** describe the SIR dynamics for scale-free and small-world networks.

### Lockdown Definition

The lock-down state is a network wide reduction of interaction probability from *p*_*i*_ to *p*_*l*_ and aims to mimic the restriction of interaction of infected nodes. The lockdown starts at *t*_0_ and is maintained for *d* time-steps. Thus *p*_*l*_ can be defined as the *mobility* of the population during the lockdown period and constraint on the social interaction. The maximum fraction of infected individuals (MFII) over the entire course of the epidemic serves as an index of lockdown efficacy. The lockdown is considered successful if the MFII stays below a specified threshold. Due to the stochastic nature of the model, results are averaged over multiple simulation runs (*n*_*t*_).

## Results

### SIR dynamics

The agent based model (ABM) can simulate the evolution of susceptible, infected and recovered populations during disease spreads [Newman, 2018]. The number of infected individuals in the population grows over time until it reaches a critical value (**Figure 1**) for both small-world and scale-free networks. Thereafter, we notice a reduction in the number of cases (infected population). Depending on the model parameters, the MFII could be as high as 1, implying that the disease has infected all individuals in the population (**Figure 1**). **Figure 1** also shows the fraction of susceptible, infected and recovered individuals at different time-points of infection spread for small-world and scale-free networks. While qualitatively the disease spreads appear to resemble the ODE-based SIR-models[Kermack and McKendrick, 1991], ABMs provide more flexibility in adding real-world complexity and simulations of interventions.

### Lockdown simulations

Lockdown scenarios could be capture by parametric control of mobility (*p*_*l*_) in ABM evolution. **Figure 2** demonstrates the effect of lockdown on disease spread (in boxed regions A, B and C). For the particular case depicted in the figure (d=30 days), we observe the existence of 3 qualitatively different scenarios. For an early lockdown (day 1), we find that the maximum fraction of infected individuals (MFII) is considerably smaller than what would have happened without a lockdown. For a slightly delayed start (day 10), infection is arrested during the lockdown period, but there is an emergence of a peak post lockdown period. Finally, if there is a late lockdown start (day 19th), the MFII reaches almost same levels as without interventions. Furthermore, we also find that a smaller duration of early lockdown cannnot prevent the MFII to reach almost the same levels of that without intervention and the existence of an optimal region (around day 9) where a relatively short lock-down is able to control the spread of the infection.

**Figure 2.**
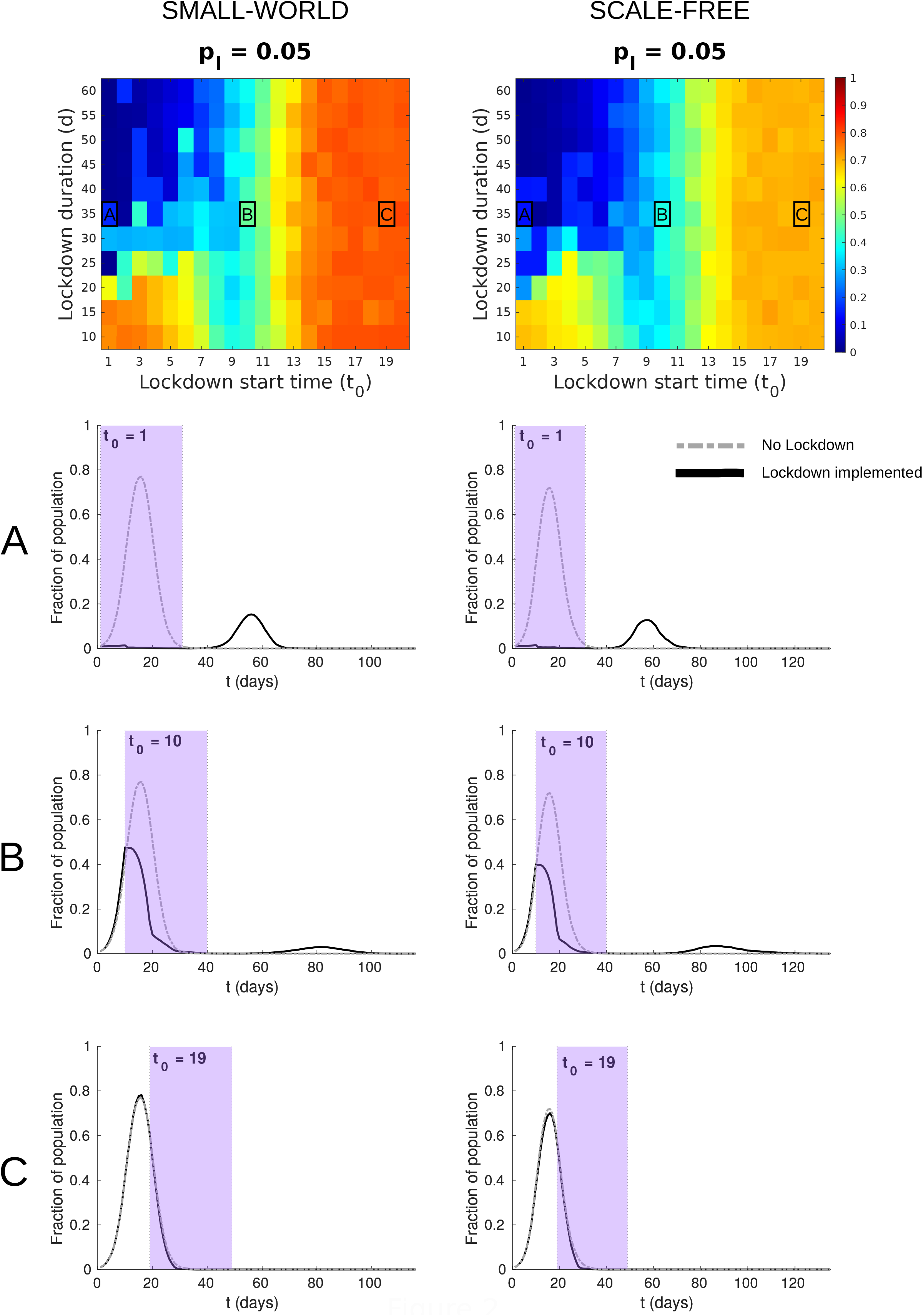
Lockdown scenarios. **a)** 3 lockdown scenarios for small-world network *N* = 1000, 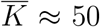 and *p*_*l*_ = 0.05. **b)** Color indicates maximum fraction of infected individuals (MFII) for different values of lockdown start time *t*_0_ and duration *d*.

### Effect of lockdown mobility

Lower values of the parameter mobility gave more favorable outcomes, i.e, reduction in MFII for a wider range of lockdown start time and duration (**Figure 3**). However, with increases in mobility (upto 0.1), we observed the size of the optimal region where a slightly delayed lockdown can achieve better results than an early lockdown. The similar pattern was observed for both small-world and scale-free networks.

**Figure 3.**
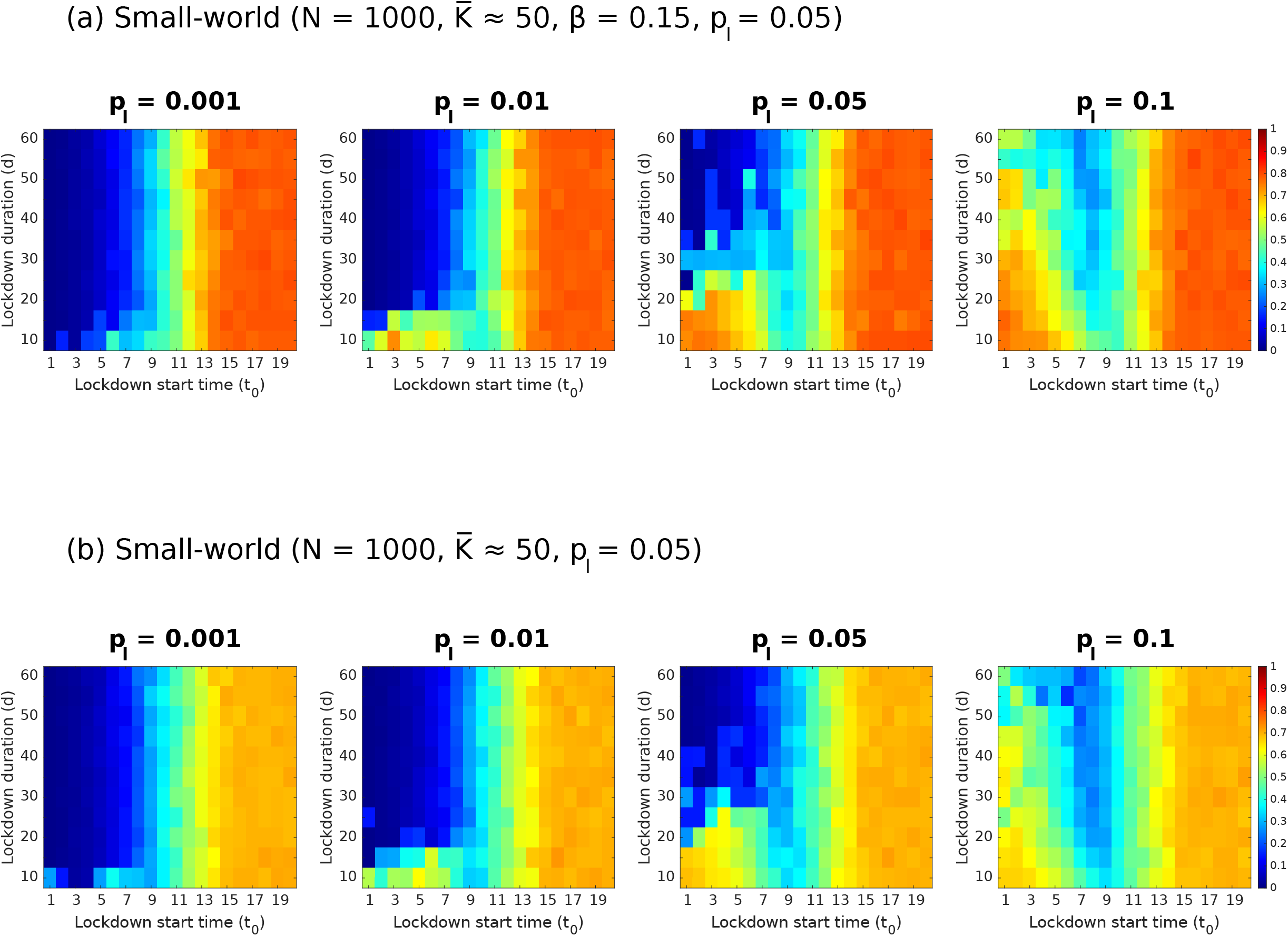
Effect of mobility during lockdown. **a)** MFII for small-world networks with various values of lockdown mobility *p*_*l*_ **b)** MFII for scale-free networks. *N* = 1000, 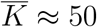, *r* = 10

### Effect of network topology and population size

**Figure 3** also suggests differences in lockdown outcomes for the two network topologies considered here, with disease spreading more aggressively on small-world topologies. An equally relevant parameter for social-networks is the overall size. Qualitatively similar results were obtained for bigger populations, as evidenced by **Figure 4** (N= 10000), however the number of active cases peaked much later(*≈* 15 days) as compared to smaller population sizes.

**Figure 4.**
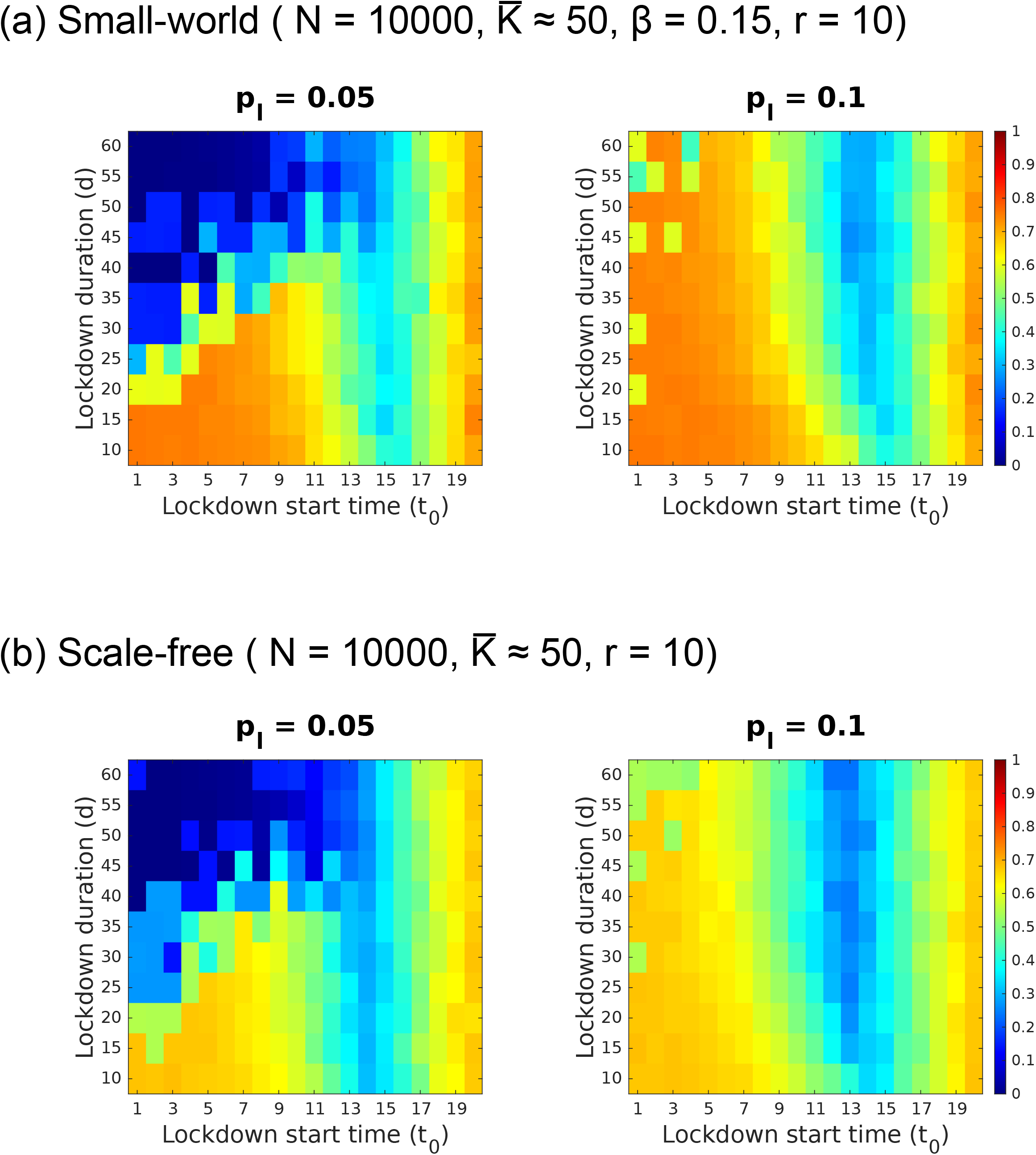
Effect of population size. MFII for *p*_*l*_ = 0.05 and *p*_*l*_ = 0.1 for small-world and **b)** scale-free networks for *N* = 10, 000.

### Effect of recovery time

The recovery time (r) of the infection has a considerable effect on both lockdown efficacy, as shown in **Figure 5**, a higher recovery time increases the overall severity of the epidemic, as is suggested by the MFII scores. Interestingly with higher recover times, the parameter space where an early lockdown with long duration would have inadvertently controlled the epidemic shrinks rapidly. Subsequently, the optimal regions where timing of a lockdown is a crcuial parameter to consider emerges as the best intervention.

**Figure 5.**
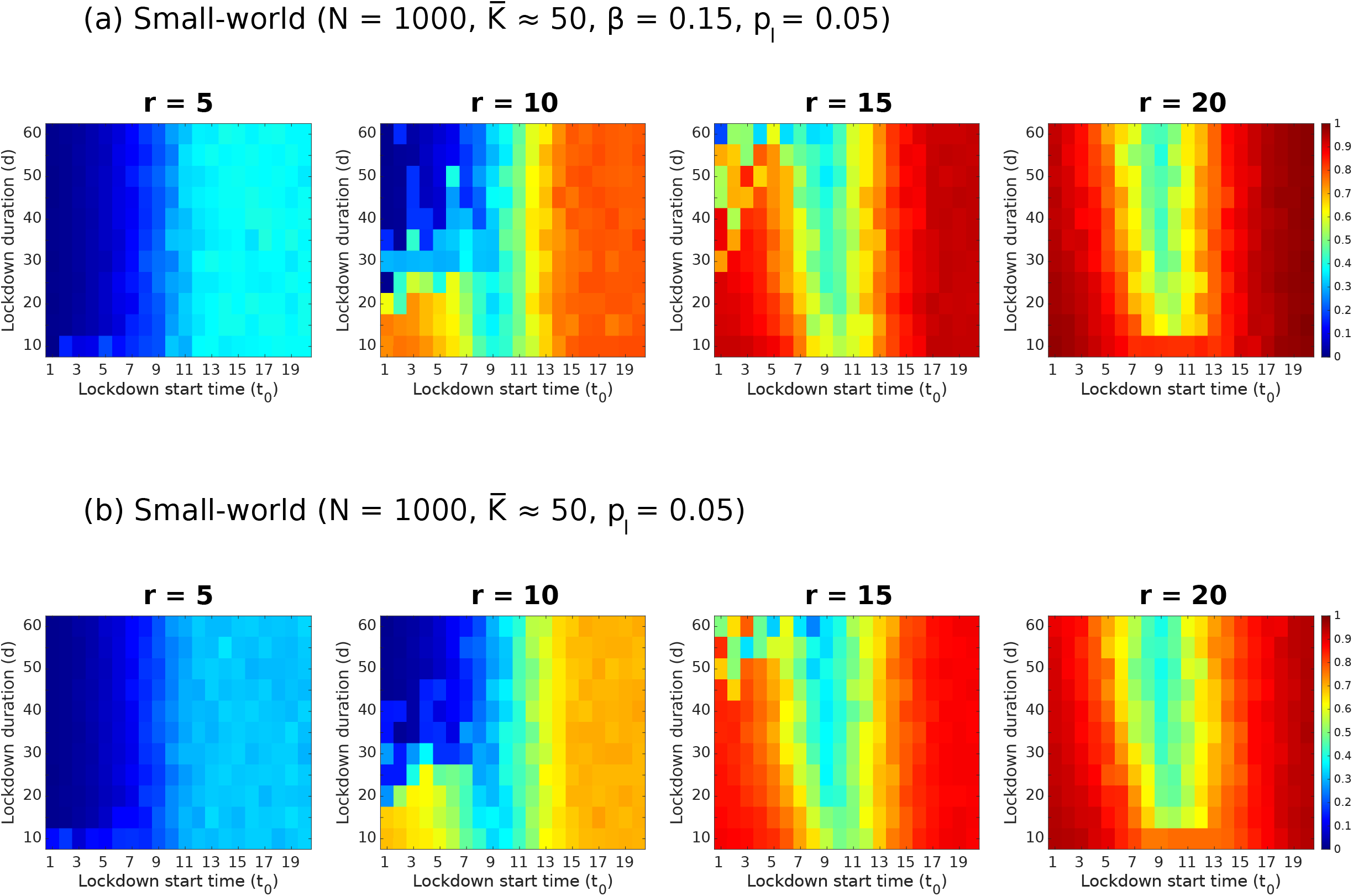
Effect of recovery time. MFII as a function of recovery time, *r* for **a)** small-world and **b)** scale-free topology.

### Optimal temporal windows for lockdown

A common theme of all our analysis is the existence of optimal temporal windows where lockdown has the maximum effect. We further evaluated how the existence of these windows change when rewiring probability is varied in small-world and scale-free networks. For small-world graphs, higher MFII values were observed for larger values of the rewiring probability (**Figure 6**). In **Figures 3, 5**, and **6**, it is interesting to note that the location of the optimal temporal window post which lockdown interventions are rendered futile remains in the vicinity of day 9. **Figures 3, 4, 5**, and **6** clearly show that a lockdown scenario is characterised by simply reducing the probability of interaction of the infected nodes with their neighbours can have a drastic effect on the severity of the epidemic on any given day. However, a lockdown event, although resulting in a low MFII, can result in more than one infection event, as illustrated in **Figure 2**. Nonetheless, since the MFII is the global upper bound on the number of individuals infected throughout the course of the epidemic, it is ideal for gauging the effectiveness of the lockdown strategy.

**Figure 6.**
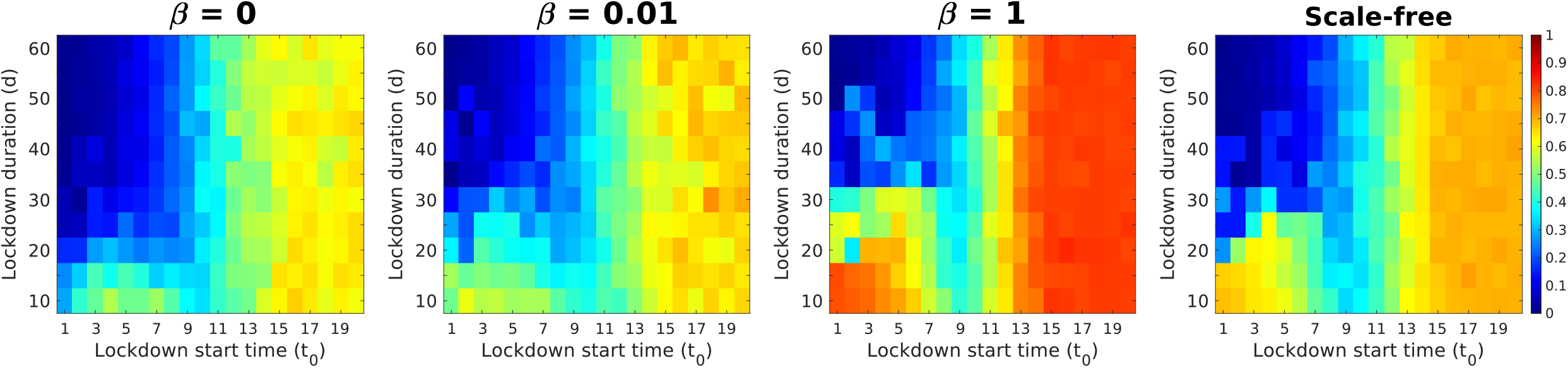
Effect of re-wiring probability. MFII for various rewiring probabilities *β* and scale-free network. 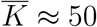.

The similar location of the optimal temporal window in Figures 2,3,5,6 indicate that this feature does not depend on the structure of the network, or on the recovery time. This effect was also observed in the larger networks, albeit around day 14/15 (Figure 4). Post this window, all lockdown strategies are rendered nugatory, no matter how long the duration of the lockdown. This alludes to the possibility of the unhindered disease crossing a certain threshold, after which it becomes impossible to arrest the infection spread using any kind of lockdown strategy. Comparison of the location of the observed temporal window with an unhindered epidemic, for both N=1000 and N=10000 suggests that this threshold lies at around *S* = 0.68 *∗N*, where S is the number of susceptible individuals.

## Discussion

We employed an agent based modeling (ABM) framework on a static network with nodes that have only one attribute - its disease state (susceptible, infected, or recovered). The parameters in this model can be broadly classified as **a)** Network parameters - node number, degree distribution, and the graph architecture, **b)**Agent parameters - mobility (interaction probability), number of interactions per day, and **c)** Disease parameters - infection probability, recovery time. Based on such a simplified parameter space we could simulate the complex dynamics of the disease spreads and study the effects of lockdown scenarios. One key result of our work is the existence of the optimal temporal windows for lockdown efficacy that can exist due to interaction of disease parameters and network topology. Second, we could demonstrate systematic differences in the severity of disease spread (as indexed by MFII) between small-world and scale-free networks. Third, our analysis reveals that early lockdowns may not be as efficacious because they limit the beneficial effects of herd-immunity and therefore risk the possibility of secondary waves of infection after the lockdown is lifted. Finally, the size of the population is found to be an important determinant to place the location of optimal lockdown windows. An important point to note is that our model is of entirely conceptual nature, but can be extended to any local community level or country-level policy making base on the knowledge of the social network topology.

### Lockdown timing

A clear outcome of our analysis presented here is that optimal lockdown windows exist and they depend sensitively on topological (network architecture) and social factors (mobility) as well as on the inherent disease parameters (e.g.,recovery time). As has been suggested elsewhere, imposing early lockdowns is no guarantee against adverse outcomes, as premature lockdowns limit the influence of herd-immunity and thereby do not preclude the possibility of aggressive post-lockdown spread. This problem is exacerbated by the leakiness inherent to any lockdown intervention. Indeed, our model is capable of predicting the possibility of *second-waves* due to ineffectual lockdowns [Xu and Li, 2020]. Also apparent is the delayed imposition of lockdowns would lead to a breakdown of essential medical services, thereby leading to high death rates. Counteracting the effect of the delay would require untenable lockdown durations. Our model predicts the existence of brief periods, close to the peak of the spread, that present windows where even moderate lockdown durations can drastically reduce the size of subsequent peaks which will be very important factors to consider for lower-to-middle income countries (LMIC).

Existence of optimal lockdown windows depends upon the choice of the quantity that we are aiming to optimize. The current analysis places a premium on the maximum fraction of infected individuals(MFII) which serves as a measure of strain on medical infrastructure at any given point of time. Indeed, recent events have demonstrated the need for *curve flattening* in order to decongest hospitals and reduce the burden on health-care professionals. However, it may be useful to frame the problem within a formal optimization framework to optimize quantities such as the basic reproduction number(*R*_0_). Similarly, it is also possible to formulate more sophisticated measures such as those derived from economic considerations [Rachel, 2020].

An important caveat of our approach is an analytical solution to optimal lockdown remains out of scope. For example, ODE-based models can also be tweaked by introducing time-dependent infection rates [Rachel, 2020, Hethcote, 2000], to genereate analytically tractable lockdown windows. However, we would like to reemphasize, a network-motivated interventions are not possible within the ODE-framework. Whether carefully selected topological modifications such as dynamic changes in network properties during lockdown period could lead to a widening or deepening of optimal lockdown windows would be an interesting avenue for future mathematical research. In a similar vein, it may be worthwhile to add even greater details to the interaction landscape such as by incorporating precise city-specific details such as population densities, age-distributions and features of transport network etc[Harsha et al., 2020].

### Role of topology

An important observation from our analysis was that the small-world networks seems to be worse affected by disease spreads in terms of MFII. However, the effect of lockdown on both the networks is almost identical. The reason for this result can be arrived at by looking at the parameters that affect disease transmission in a network in relation to the lockdown strategy employed. In this study, the lockdown is an intervention which alters the usual course of the epidemic by abruptly reducing the probability of infection spread by restricting the interactions of infected individuals while not affecting the change in the structure of neighbourhoods in both types of networks. Thus, the lockdown can be viewed as a control of dynamics on a given set of structural constraint but not directly affecting the interaction properties between nodes. The effect of reducing interaction probability to various values of *p*_*i*_ is clearly shown in Figure 3. The default value of *p*_*i*_ is 0.5, which implies that an infected node has a fifty percent chance of interacting with a randomly chosen neighbour. Naturally, reducing this value of *p*_*i*_ to the extreme value of 0 would imply that an infected node interacts with absolutely no neighbour throughout the lockdown. In this case, if the duration of the lockdown is such that it exceeds the recovery time of the disease, the infection would die out. Such extreme values of *p*_*i*_ seem implausible in a real-world setting, and so we assume non-zero values of *p*_*l*_, or *“leaky lockdowns”*. The lowest value of *p*_*i*_ used in this study is 0.001, which is low enough to render even a 10-day lockdown highly efficacious, as long as it is implemented early enough. As *p*_*l*_ is increased, we see that lockdowns of small duration are not efficacious even if implemented at an early stage.

### Population Size Effects

Simulation results indicate the peak in active cases occurs later for larger as compared to smaller populations for the same number of local connections. This is a crucial consideration while comparing lockdown timings between countries with different population sizes. Interestingly, the optimal lockdown timing seems to shift between N = 1000 and N= 10,000 **(Figure 4)**. This change could be explained by observing that the mean degree for both N = 1000 and N = 10,000 is kept at 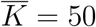 and therefore, the spreading dynamics is delayed for the case of larger population. One could expect 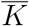 to scale with population size, but research has suggested that the local contact rate remains constant across larger and smaller populations[Hethcote, 2000]. How each one of these factors can be applied to urban and rural situations and can be relevant for community/ nation specific policy making remains scope for future research.

## Data Availability

Codes will be available on request

## Acknowledgements

This work was supported by NBRC core funds.

## Abbreviations

ABM: Agent Based Modelling
SIR: Susceptible, Infected, Recovered/Removed
MFII: Maximum Fraction of Infected Individuals
ODE: Ordinary differential equation

## Figure Legends

*N*_*I*_ = 10 and *n*_*t*_ = 5 for all the cases unless otherwise stated.

